# Functional germline variants in DNA damage repair pathways are associated with altered survival in adults with glioma treated with temozolomide

**DOI:** 10.1101/2023.10.13.23296963

**Authors:** Geno Guerra, George Wendt, Lucie McCoy, Helen M. Hansen, Linda Kachuri, Annette M. Molinaro, Terri Rice, Victoria Guan, Lianne Capistrano, Allison Hsieh, Veruna Kalsi, Jaimie Sallee, Jennie W. Taylor, Jennifer L. Clarke, Eduardo Rodriguez Almaraz, John K. Wiencke, Jeanette E. Eckel-Passow, Robert B. Jenkins, Margaret Wrensch, Stephen S. Francis

**Author notes:** **Corresponding Author** Stephen S. Francis, PhD, Associate Professor, Division of Neuro and Molecular Epidemiology, Department of Neurological Surgery, University of California San Francisco, 1450 3rd St, San Francisco, CA 94158.

## Abstract

**Background:** Temozolomide (TMZ) treatment has demonstrated, but variable, impact on glioma prognosis. This study examines associations of survival with DNA repair gene germline polymorphisms among glioma patients who did and did not have TMZ treatment. Identifying genetic markers which sensitize tumor cells to TMZ could personalize therapy and improve outcomes.

**Methods:** We evaluated TMZ-related survival associations of pathogenic germline SNPs and genetically predicted transcript levels within 34 DNA repair genes among 1504 glioma patients from the UCSF Adult Glioma Study and Mayo Clinic whose diagnoses spanned pre- and post-TMZ eras within the major known glioma prognostic molecular subtypes.

**Results:** Among those who received TMZ, 5 SNPs were associated with overall survival, but not in those who did not receive TMZ. Only rs2308321-G, in *MGMT*, was associated with decreased survival (HR=1.21, p=0.019) for all glioma subtypes. Rs73191162-T (near *UNG*), rs13076508-C (near *PARP3*), rs7840433-A (near *NEIL2*), and rs3130618-A (near *MSH5*) were only associated with survival and TMZ treatment for certain subtypes, suggesting subtype-specific germline chemo-sensitization.

Genetically predicted elevated compared to normal brain expression of *PNKP* was associated with dramatically worse survival for TMZ-treated patients with *IDH*-mutant and 1p/19q non-codeleted gliomas (p=0.015). Similarly, *NEIL2* and *TDG* expressions were associated with altered TMZ-related survival only among certain subtypes.

**Conclusions:** Functional germline alterations within DNA repair genes were associated with TMZ sensitivity, measured by overall survival, among adults with glioma, these variants should be evaluated in prospective analyses and functional studies.

**Key points:**

- We observed SNPs associated with glioma survival, specific to cases receiving TMZ
- An *MGMT* variant may reduce glioma survival indirectly through myelosuppression
- Decreased genetic *PNKP* expression in the brain may sensitize cells to TMZ

**Importance of the study:** The introduction of temozolomide (TMZ) as a part of standard-of-care in the treatment of gliomas marked the last notable increase in patient survival. However, the effectiveness of TMZ is not universal, and can result in serious complications. The mechanism of action behind the drug is the introduction of damaging methyl groups across the tumor genome and leveraging of DNA damage repair (DDR) mechanisms to signal programmed cell death. Previous literature has identified that defects in DDR mechanisms can alter TMZ sensitivity. Using a unique dataset that spans the pre- and post-TMZ eras, we demonstrate that germline variation in DDR-related genes may have significant impact on overall survival for patients treated with TMZ, with no effects observed in the pre-TMZ era. This suggests that germline variants in these DDR genes could be used to personalize TMZ therapy to improve patient survival.

## Introduction

Gliomas are a histologically and molecularly diverse group of highly fatal cancers originating from brain glial cells or their precursors. Overall, gliomas have an incidence rate of ∼6 per 100,000 persons^1^. Glioma prognoses can be well stratified by the identification of somatic markers within the primary tumor, the most prominent features being mutations to isocitrate dehydrogenase 1 and 2 (*IDH*) genes, and codeletion of the 1p and 19q chromosomal arms (1p/19q)^2^. The most aggressive subtype, Glioblastoma, *IDH*-wildtype (GBM), has a median overall survival rate of roughly 1.2 years^3^. The last notable increase in glioma patient survival coincided with introducing the chemotherapy agent temozolomide as part of standard-of-care in 2005^4^.

Temozolomide (TMZ), is an alkylating chemotherapy that induces DNA damage in tumor cells through the methylation of purine bases, creating genomic instability upon replication, and is used across glioma subtypes given its blood-brain barrier penetrability. Although O^6^-methylguanine-DNA-methyltransferase (*MGMT*) promoter methylation is known to increase TMZ sensitivity in gliomas, other molecular pathways, mismatch repair (MMR), and base-excision repair (BER) pathways also play essential roles in inducing DNA damage from TMZ. These pathways involve the identification and removal of TMZ-induced alkylated bases, leading to either cell death, as intended by the drug, or possibly cell repair, resulting in decreased TMZ efficacy^5^. From its initial introduction in 1999, to its widespread adoption in 2005^4^, TMZ has now become standard-of-care in treating *IDH*-wildtype glioblastomas, alongside maximum safe surgical resection and adjuvant radiation therapy. Some patients’ tumors, however, are resistant to treatment^6^.

TMZ has also been used to treat *IDH*-mutant gliomas, but its utility has been more controversial. Treatment with TMZ has been associated with possible somatic tumor hypermutation leading to recurrence and high-grade transformation in patients with primary grade 2/3 *IDH*-mutant glioma. These tumors exhibit thousands of new coding mutations, that generate a distinctive TMZ-induced hypermutation signature^7,8^, including somatic defects in DDR genes. Cells lacking MMR function cannot recognize alkylated bases, leading to unrepaired DNA alterations distributed throughout the genome^9^. One study observed that amongst 82 low grade *IDH*-mutant gliomas (who previously received TMZ treatment) undergoing re-operation for recurrent transformed tumors, roughly 57% of cases experienced TMZ-induced hypermutation at recurrence, which was subsequently associated with shorter patient survival^9^. Although there is still a clinical knowledge gap in characterizing if genetic markers can predispose patients to TMZ resistance and/or TMZ-induced hypermutation, recent findings reveal that somatic *MGMT* promoter methylation levels in newly diagnosed grade II/III gliomas are predictive of hypermutation at recurrence and may serve as a prospective biomarker to inform clinical decision making^10^. However, comprehensive explanations for resistance to TMZ and variability in tumor hypermutation in the absence of *MGMT* promoter methylation remain unknown. One possible avenue could be the influence of germline polymorphisms in *MGMT*, MMR and BER pathway genes in TMZ treated patients.

Many germline polymorphisms, particularly SNPs in/near genes involved in DDR pathways, have been associated with altered genetic predispositions to cancers, such as glioma, colorectal, lung, breast, and others^11–14^. Since the response to TMZ is variable in patients with glioma, we hypothesized that polymorphisms in *MGMT* and BER/MMR genes may be associated with altered TMZ-specific glioma survival outcomes as a result of altered/impaired drug function. To test this hypothesis, we conducted both SNP and transcript association studies of germline alterations across 33 BER/MMR-associated genes and *MGMT* with overall survival in 1504 adults with glioma and known presence or absence of TMZ usage. The longevity of the cohort collection utilized in this study provided a unique opportunity to study the effects of TMZ usage without the selection bias. The cohort of patients that comprise this study spans from 1991-2014 providing a large number of glioma patients prior to and after the introduction of TMZ to the US market.

## Materials and Methods

### Study Populations

Our study included 1504 patients with newly diagnosed glioma collected between 1989 and 2014 from the University of California, San Francisco or the Mayo Clinic with available genotyping, molecular subtyping (*IDH* mutation, 1p/19q codeletion status), tumor grade, treatment, demographic, and survival information. 994 cases were genotyped on the Illumina OncoArray, as previously described^15–18^. 510 cases were genotyped on the Illumina HumanHap370duo panel (i370)^17^. These datasets data quality control, imputation, and sample selection details have been previously described^19^.

For subtype-specific analyses, samples were separated into major subtypes using presence/absence of somatic *IDH* mutation and 1p/19q chromosomal arm co-deletion^3^. Recorded temozolomide usage (yes/no) during first-course treatment (treatment prior to disease progression/recurrence) was required for all cases for inclusion. Treatment information was previously abstracted from medical records or the California Cancer Registry, where applicable.

### Study Genes

Thirty-three genes were selected in the BER and MMR DNA repair pathways as well as *MGMT* (for a total of 34 considered genes) utilizing information collected by Wood et al. at MD Anderson Cancer center^20^. The gene list included 22 BER genes, and 11 MMR genes, irrespective of previous literature linking their function to TMZ chemosensitivity (list of 34 genes available in **Figure 1**).

**Figure 1:**
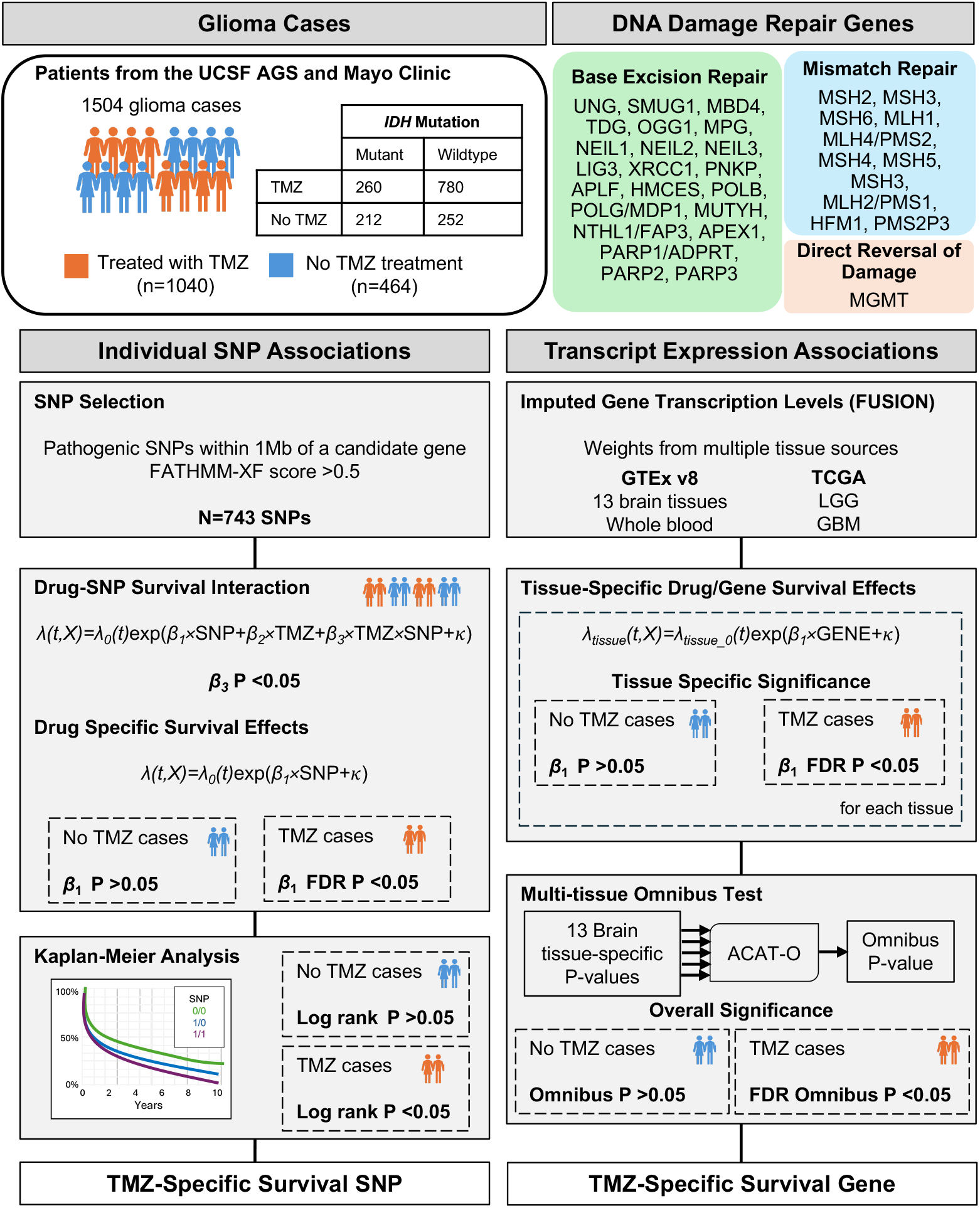
Overview of study design. A visual outline of the sample set of glioma cases included, DNA damage repair genes selected, and pipeline overviews for both the individual SNP analyses and genetic transcript expression analyses.

### Individual SNP Selection

For single variant analyses, all single nucleotide variants (SNVs) within a 1Mb window of the 34 included DNA repair genes (defined as 500kb upstream or downstream of the gene boundary) were considered for inclusion in the study. We first filtered for SNVs available (after imputation) on both arrays (Oncoarray and i370), and then kept only those SNVs with population minor allele frequency (MAF) >0.01 (amongst European populations CEU, TSI, FIN, GBR, and IBS from the 1000 Genomes Project^21^). CADD (v1.3)^22^, and FATHMM-XF^23^ were used to annotate and predict possible functional/deleterious variants. SNVs with FATHMM-XF coding or non-coding scores >0.5 (predicted to be pathogenic variants) were retained for individual SNP analyses.

### Single SNP association analysis

To evaluate the relationship between predicted pathogenic variants near the 34 genes of interest and TMZ-specific survival, we utilized a pipeline adapted from Rong et al.^24^ (**Figure 1**). Subtype specific Cox Proportional Hazard (Cox PH) regression models with included SNP-TMZ interaction term were fit using gwasurvivr^25^ with further adjustments for age at diagnosis, sex, tumor grade, *IDH* status, 1p/19q codeletion, radiotherapy, diagnosis year, data source (UCSF AGS or Mayo Clinic), and the top 10 genetic PCs, where appropriate:

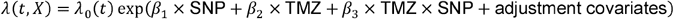

SNPs were modeled as continuous dosage values. TMZ usage was binary yes/no based on medical records indicating TMZ given in first-course treatment. Follow-up time was defined as the difference between date of surgery-confirming diagnoses to either date of death or censored last known contact. To account for array-specific biases, all Cox survival analyses were stratified by genotyping array and effects were combined using a fixed effects meta-analysis.

For nominally significant SNP-TMZ interactions (p<0.05 for *β*_3_), Cox PH models were fit separately for the TMZ and no-TMZ groups, adjusting for identical covariates. SNPs with significant survival associations in the no-TMZ group (p<0.05) were dropped from analysis. Remaining SNPs with significant survival associations in the TMZ group (FDR adjusted p <0.05) were retained.

Kaplan-Meier (KM) survival analyses were used as a final step to confirm if the candidate SNPs were explicitly associated with altered survival outcomes only in those who received TMZ and that they had no survival association in the no-TMZ group. SNPs with a KM p-value p<0.05 in the no-TMZ group were discarded. Remaining SNPs with a KM p-value p<0.05 in the temozolomide users’ group were ultimately determined to be significantly associated with TMZ-mediated glioma survival (**Figure 1**).

### Transcriptome association analysis

Gene transcription levels for the 34 genes of interest were imputed for each included sample using the FUSION approach^26^. We utilized pre-computed SNP weights calculated using tissue-specific samples from GTEx v8 and TCGA, as made publicly available on the FUSION website (http://gusevlab.org/projects/fusion). Scores were calculated from GTEx using 13 different brain location-specific, and whole blood specific weights. Scores using tumor sample expression data were calculated from glioma tissue samples from the glioblastoma (GBM) and low-grade glioma (LGG) TCGA projects. Genetically imputed gene expression scores were analyzed as continuous variables using a Cox PH regression framework. Models were adjusted for age at diagnosis, sex, tumor grade, *IDH* status, 1p/19q codeletion, whether patients received radiotherapy, diagnosis year, and the top 10 genetic PCs. Models were stratified by genotyping array and combined using a fixed-effects meta-analysis. Hazard ratios were estimated separately for cases treated with or without reported TMZ treatment.

We utilized ACAT-O^27^, a robust omnibus aggregated Cauchy association test, to combine p-values across the 13 available GTEx brain tissues for each gene association. Significant effects in the no-TMZ group (*P*_ACAT-O_<0.05) were determined to be not drug specific and were discarded. Remaining genes with significant effect in the TMZ group (FDR-adjusted *P*_ACAT-O_<0.05) were determined to be drug specific. Genes with FDR *P*_ACAT-O_<0.1 are also reported as suggestively associated with TMZ-related survival (**Figure 1**).

## Results

### Characteristics of the included samples

A total of 1504 patient samples with glioma (472 *IDH*-mutant: 168 1p/19q codel, 304 non-codel, and 1032 *IDH*-wildtype glioblastoma) with known TMZ treatment (present or absent) during first round of treatment were included for analysis. Clinical and demographic information of the included glioma cases stratified by known TMZ usage is summarized in **Table 1**. 75.6% of cases with an *IDH*-wildtype glioblastoma received TMZ treatment, compared to only 55.0% of cases with an *IDH*-mutated glioma (**Supplemental Table S1**).

**Table 1:** Clinical and molecular summary of included adults with glioma by known temozolomide usage.

|  | Received TMZ | No TMZ |
| --- | --- | --- |
| Number of cases | 1040 | 464 |
| Median age at diagnosis (IQR) | 54 (18) | 49 (24) |
| Sex (% Male) | 63.65% | 58.62% |
| Genotyping array |  |  |
| Oncoarray | 68.46% | 60.78% |
| i370 | 31.54% | 39.22% |
| Data source |  |  |
| UCSF Adult Glioma Study | 77.02% | 72.20% |
| Mayo Clinic | 22.98% | 27.80% |
| *IDH* |  |  |
| Wildtype | 75.00% | 54.31% |
| Mutant | 25.00% | 45.69% |
| 1p/19q |  |  |
| Codeleted | 8.94% | 17.46% |
| Non-codeleted | 81.35% | 75.43% |
| NA | 9.71% | 7.11% |
| *TERT* |  |  |
| Wildtype | 23.94% | 32.33% |
| Mutant | 59.62% | 47.84% |
| NA | 16.44% | 19.83% |
| Grade |  |  |
| 2 | 11.73% | 37.07% |
| 3 | 19.33% | 19.18% |
| 4 | 68.94% | 43.75% |
| Histology |  |  |
| Astrocytoma | 18.46% | 24.35% |
| Oligodendroglioma | 7.50% | 18.53% |
| Glioblastoma* | 68.56% | 43.32% |
| Oligoastrocytoma | 5.48% | 13.79% |
| Diagnosis Year |  |  |
| Median (min, max, IQR) | 2007 ('98, '14, 6) | 2002 ('91,'14, 9) |
| % Censored (no known death event) | 16.35% | 23.92% |
| Chemotherapy (% known Chemo) | 100% | 34.91% |
| Radiation (% known RT) | 90.67% | 71.12% |

\*Group contains *IDH* wildtype and mutant. World Health Organization 2021 guidelines now classify *IDH* mutant Glioblastoma as Grade 4 Astrocytoma

### Individual DNA repair variants and TMZ specific survival

We individually examined the association between 743 SNPs with predicted pathogenicity (defined by FATHMM-XF score >0.5) and glioma survival time for cases with and without known TMZ treatment. All analyses were conducted for glioma overall and stratified by presence of *IDH* mutation and 1p/19q co-deletion status (**Table 2, Figure 2, Supplementary File 1**).

**Table 2:** Genetic variants near DNA damage repair genes associated with temozolomide-specific survival amongst glioma molecular subtypes.

| Nearby Gene | rsid | Chr | Pos | Ref Allele | Effect Allele | Effect Allele Freq. | SNP consequence | CADD PHRED | FATHMM score | FATHMM classification | Interaction P-value | TMZ cox PH model |  |  |  | No-TMZ cox PH model |  |  |
| --- | --- | --- | --- | --- | --- | --- | --- | --- | --- | --- | --- | --- | --- | --- | --- | --- | --- | --- |
|  |  |  |  |  |  |  |  |  |  |  |  | HR (95% CI) | Cox P | Cox P (FDR adjusted)* | KM P | HR (95% CI) | Cox P | KM P |
| All glioma |  |  |  |  |  |  |  |  |  |  |  |  |  |  |  |  |  |  |
| MGMT | rs2308321 | 10 | 129766800 | A | G | 0.12 | missense | 8.74 | 0.63 | pathogenic | 0.028 | 1.21 (1.05-1.4) | 0.0091 | 0.019 | 0.011 | 0.98 (0.77-1.25) | 0.87 | 0.34 |
| IDH-wildtype glioblastoma |  |  |  |  |  |  |  |  |  |  |  |  |  |  |  |  |  |  |
| UNG | rs73191162 | 12 | 109222793 | C | T | 0.037 | splice, intron | 0.63 | 0.56 | pathogenic | 0.042 | 1.73 (1.31-2.29) | 0.0001 | 0.0004 | 0.0007 | 0.91 (0.55-1.51) | 0.71 | 0.56 |
| IDH-mutant glioma |  |  |  |  |  |  |  |  |  |  |  |  |  |  |  |  |  |  |
| PARP3 | rs13076508 | 3 | 52373789 | T | C | 0.050 | non-coding exon | 15.04 | 0.84 | pathogenic | 0.029 | 0.4 (0.18-0.88) | 0.023 | 0.026 | 0.041 | 1.16 (0.65-2.06) | 0.62 | 0.93 |
| IDH-mutant 1p/19q codeleted glioma |  |  |  |  |  |  |  |  |  |  |  |  |  |  |  |  |  |  |
| NEIL2 | rs7840433 | 8 | 11564661 | G | A | 0.38 | downstream | 1.01 | 0.92 | pathogenic | 0.037 | 2.33 (1.23-4.35) | 0.0087 | 0.023 | 0.023 | 1.16 (0.65-2.08) | 0.61 | 0.72 |
| IDH-mutant 1p/19q non-codeleted glioma |  |  |  |  |  |  |  |  |  |  |  |  |  |  |  |  |  |  |
| MSH5 | rs3130618 | 6 | 31664357 | C | A | 0.17 | missense | 26.80 | 0.61 | pathogenic | 0.023 | 0.58 (0.38-0.89) | 0.012 | 0.020 | 0.016 | 1.12 (0.67-1.87) | 0.67 | 0.37 |

L2 \*: FDR adjusted for number of SNPs with significant interaction P and non-significant no-TMZ cox P.
L3 KM P: Unadjusted Kaplan-Meier log rank p-value

**Figure 2:**
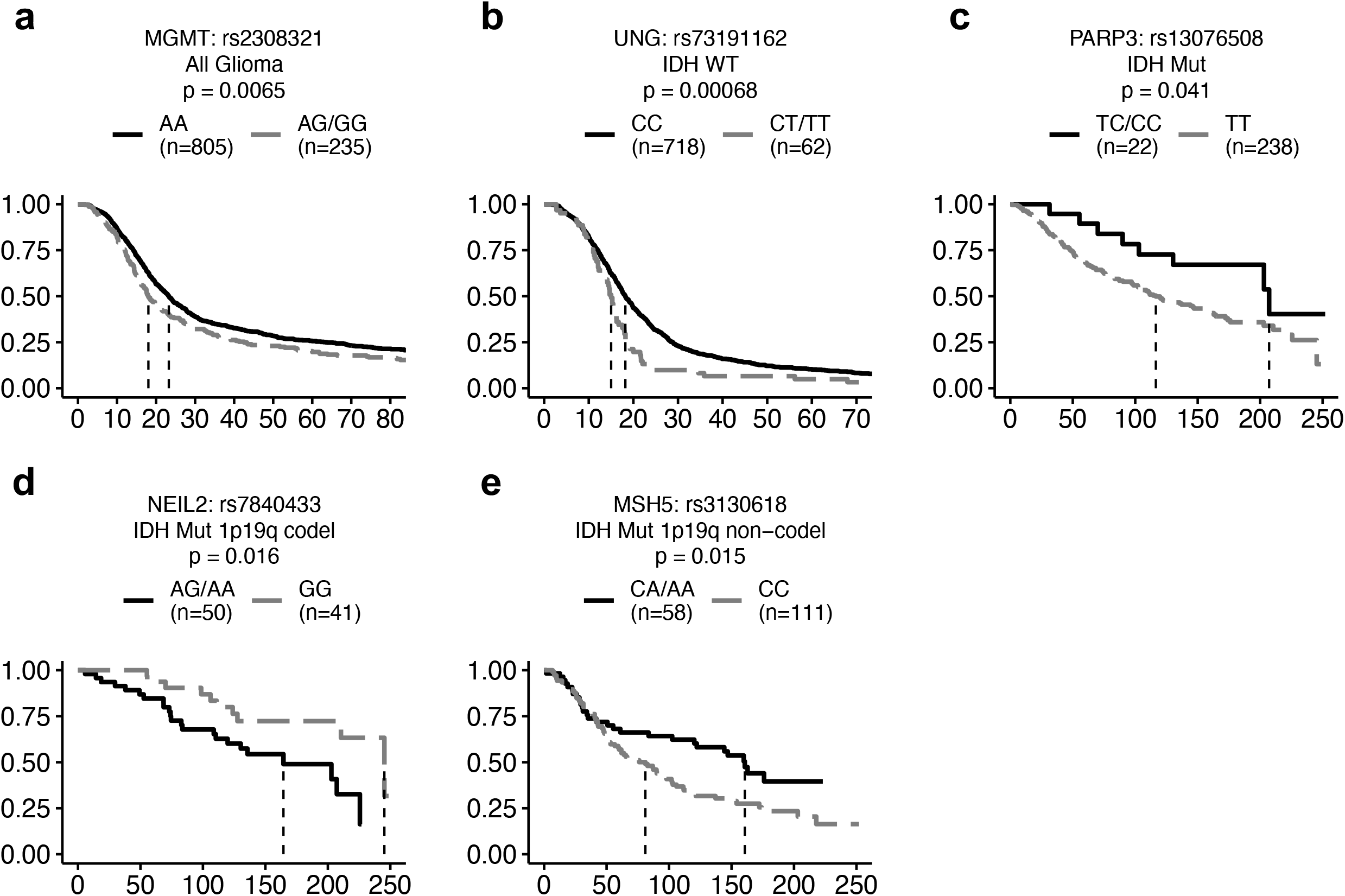
Kaplan-Meier plots visualizing the survival times associated with germline polymorphisms in subtype-specific glioma patients treated with temozolomide. Plots demonstrate the survival trajectories related to different polymorphisms of cases treated with the drug temozolomide, for a specific glioma subtype, as indicated. The x-axes represent time measured in months post-diagnosis. Dashed vertical lines represent median survival times. Log rank p-values measuring significance of statistical differences between curves are as indicated in each plot. Included number of cases with genotype of interest are indicated in parentheses.

We found that rs2308321-G, a missense variant of *MGMT*, was associated with decreased survival in glioma (overall) cases treated with TMZ (HR=1.21, 95% CI: 1.05-1.40, FDR p=0.0186), while having no detectable effect on cases without TMZ treatment (HR=0.98, 95% CI: 0.77-1.25, p=0.8717). Amongst the TMZ treated group, cases with the G polymorphism (1 or 2 copies) showed a median 5.5-month shorter survival than cases with the AA genotype (p=0.0065, **Figure 2a**). Median survival decrease of 1.2 months was not significant in the no-TMZ group (p=0.08717).

*IDH*-mutant cases carrying rs13076508-C, a SNV near *PARP3* on chromosome 3, had significantly improved survival over those without the polymorphism, amongst those treated with TMZ (HR=0.40, 95% CI: 0.18-0.88, FDR p=0.0263), with a median 90.7 months increase in survival time (p=0.041, **Figure 2c**). We observed no difference among *IDH*-mutant cases without TMZ usage based on this genotype (HR=1.16, 95% CI: 0.65-2.06, p=0.6174), and no difference in survival curves (p=0.9322).

For *IDH*-mutant 1p/19q-codeleted gliomas, the subtype with the generally best prognosis, we found that amongst TMZ-treated cases, rs7840433-A (near *NEIL2*) was significantly associated with decreased survival times (HR=2.33, 95% CI: 1.23-4.35, FDR p=0.0234), with carriers of the variant having a median 81 months decreased survival time (p=0.0229, **Figure 2d**). Cases without TMZ treatment had no significant difference in survival based on observed genotype (HR=1.16, 95% CI: 0.65-2.08, p=0.6083).

We found that rs3130618-A, in chromosome 6 near *MSH5*, was associated with increased survival among TMZ-treated *IDH*-mutant and 1p/19q non-codeleted (HR=0.58, 95% CI: 0.38-0.89, FDR p=0.0202), with A-allele carriers having a median 79.6-month improved survival time over TMZ-treated cases and the CC genotype (p=0.016, **Figure 2e**). Amongst the 135 cases not treated with TMZ, we observed no difference in survival outcomes (HR=1.12, 95% CI: 0.67-1.87, p=0.6707).

For *IDH*-wildtype glioblastoma cases, we observed that rs73191162-T, near *UNG*, was significantly associated with TMZ-treated survival (HR=1.73, 95% CI: 1.31-2.29, FDR p=0.0004), and not significant amongst no-TMZ cases (HR=0.91, 95% CI: 0.55-1.51, p=0.7127). Amongst TMZ-treated *IDH*-wildtype cases, those with any copy of the T allele had a median 3.2 month shorter survival time than those with the common CC genotype (**Figure 2b**). Cases with *IDH*-wildtype glioblastomas not treated with TMZ had no significant difference in survival based on genotype (p=0.5616).

### Functional characterization of findings

All SNPs included in the analysis had FATHMM-XF scores >0.5 (coding or non-coding), meaning they were predicted to be deleterious by the algorithm. Of the 5 significant SNPs, 2 were also predicted to be in the top 10% of deleterious polymorphisms in the genome (rs13076508 and rs3130618), based on CADD scores >10, with rs3130618 (near *MSH5*) predicted to be amongst the top 1% (CADD >20). The former SNP, rs13076508, located near *PARP3* on chromosome 3 is a significant eQTL for genes *TLR9* and *BAP1*, as well as a significant sQTL for *NISCH* and *DNAH1*, all within brain tissues (GTEx V8, *data not shown*). The latter SNP, rs3130618, is a missense SNP located in the *GPANK1* gene (exon 3) within the HLA region of chromosome 6, which results in an amino acid change from Arginine to Leucine. It is also associated with decreased *MSH5* expression within the cerebellum (p=0.0196, GTEx V8). The *MGMT* missense variant, rs2308321-G, induces a change in the amino acid sequence of the resulting *MGMT* protein (Ile143Val), which has been suggested to increase protein expression, compared to the wildtype^28^.

### DNA repair transcriptome survival association analysis

We considered genetically inferred expression in all 13 available brain tissues and whole blood from GTEx (v8) and expression within tumor samples from the of lower grade glioma and glioblastoma TCGA projects (TCGA-LGG and TCGA-GBM) (Full results in **Supplementary File 2**). Survival association concordance across brain tissues was summarized using the omnibus method, ACAT-O, for each glioma subtype (**Figure 3a**). Glioma subtype specific Cox analyses of genetically imputed expression levels identified 6 genes as significantly associated with TMZ-specific survival time in at least one tissue (Single-tissue FDR-adjusted p<0.05, **Table 3**). A further 11 suggestive gene associations (Single-tissue FDR-adjusted p<0.1) in at least one glioma subtype/tissue pair are also reported (**Supplementary Table S2**). Using the ACAT-O omnibus test across all 13 brain tissues, *PNKP* was significant (FDR P_ACAT-O_<0.05), and *NEIL2* suggestive (FDR P_ACAT-O_<0.1) with subtype specific survival after TMZ treatment.

**Table 3:** Brain tissue-specific genetically imputed transcript levels of DNA repair genes are associated with survival amongst glioma cases treated with temozolomide.

| Gene | Chr | Brain Site (GTEx) | HR (95% CI) | Cox P-value<br>(FDR adjusted) |
| --- | --- | --- | --- | --- |
| **IDH-wildtype glioblastoma** |  |  |  |  |
| *TDG* | 12 | Caudate basal ganglia | 0.88 (0.82-0.95) | 0.029 |
| **IDH-mutant glioma** |  |  |  |  |
| *HMCES* | 3 | Cerebellum | 0.74 (0.61-0.89) | 0.044 |
| *MBD4* | 3 | Frontal cortex BA9 | 1.36 (1.12-1.65) | 0.042 |
| *PNKP* | 19 | Substantia nigra | 1.38 (1.14-1.68) | 0.023 |
| **IDH-mutant 1p/19q codeleted glioma** |  |  |  |  |
| *NEIL2* | 8 | Putamen basal ganglia | 2.43 (1.44-4.10) | 0.015 |
| *NEIL2* | 8 | Spinal cord cervical c-1 | 2.25 (1.36-3.75) | 0.038 |
| *POLG* | 15 | Putamen basal ganglia | 1.89 (1.22-2.94) | 0.037 |
| **IDH-mutant 1p/1q non-codeleted glioma** |  |  |  |  |
| *PNKP* | 19 | Substantia nigra | 1.65 (1.30-2.10) | 0.0009 |

**Figure 3:**
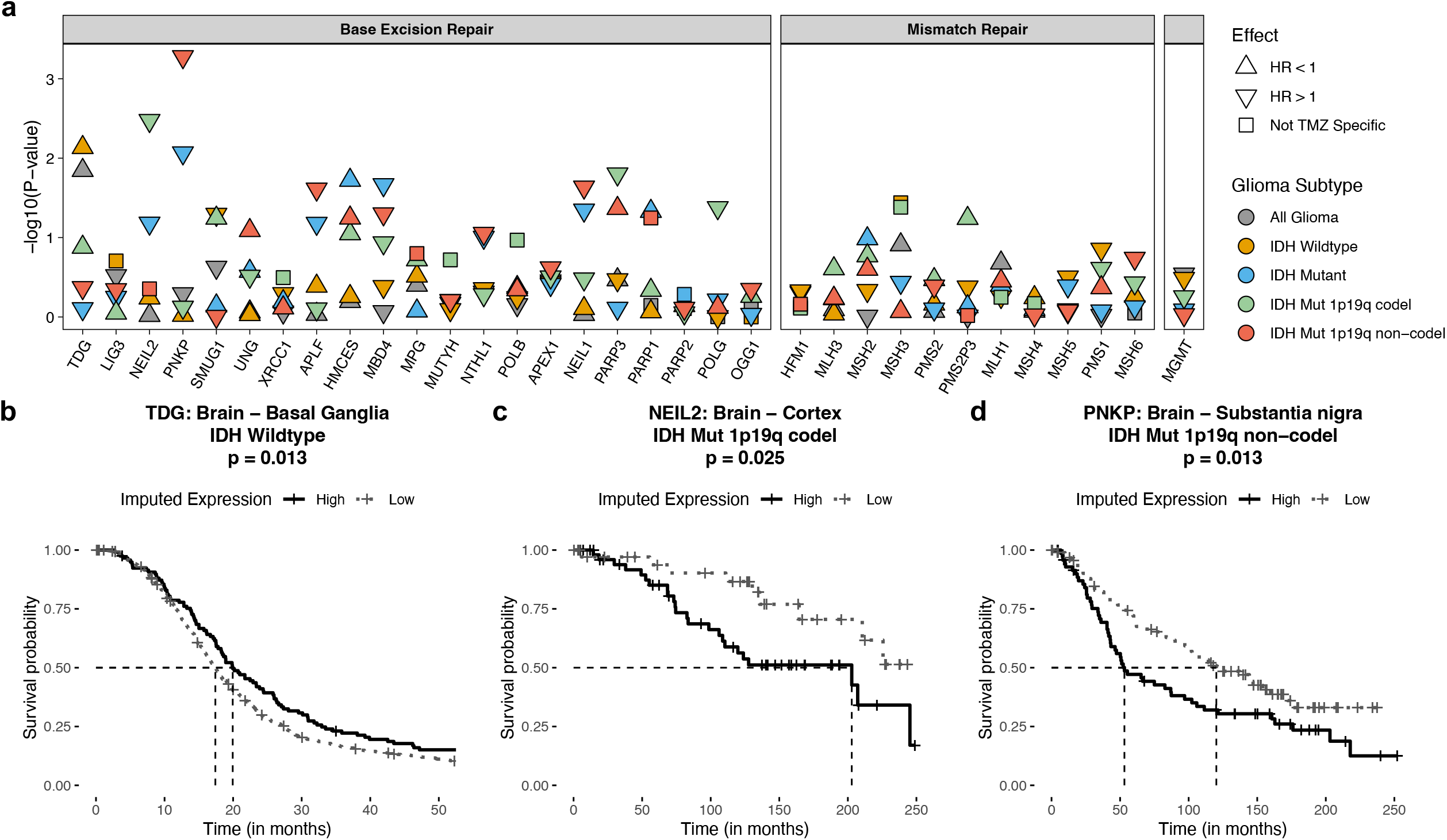
Genetically imputed DNA repair gene transcript level associations with subtype specific glioma survival, for patients who were treated with temozolomide. **a)** Nominal ACAT-O omnibus p-values for cases treated with temozolomide. Colors indicate glioma subtype tested. Shape indicates direction of effect for increasing expression. Results marked by a square indicate ACAT-O p-value <0.05 in the complementary no-TMZ group. **b)** Elevated genetically imputed transcript levels of the gene *TDG* using weights from the GTEx brain (caudate basal ganglia) show prognostic benefit to *IDH* wildtype glioma cases treated with TMZ. ‘Low’ is genetically imputed level of zero, ‘High’ is expression >0. **c)** Elevated imputed expression of *NEIL2* (using GTEx brain cortex weights) is associated with decreased survival for cases with *IDH* mutant 1p19q codeleted gliomas treated with TMZ. ‘Low’ is genetically imputed level of zero, ‘High’ is expression >0. **d)** Elevated imputed expression of the gene *PNKP* is associated with significantly decreased survival times in cases with *IDH* mutant 1p19q non-codeleted gliomas treated with TMZ. Using GTEx brain-substantia nigra weights. ‘Low’ represents the bottom 40% of expressors, ‘High’ represents the top 60%.

No genes were significantly associated with overall glioma survival differences specifically for TMZ-treated cases, rather we observed *IDH* subtype-specific associations (**Figure 3**).

The analysis identified most associations being specific to *IDH*-mutant response to TMZ. Increased genetically predicted expression levels of *PNKP* (HR=1.38, 95% CI: 1.14-1.68, Brain – Substantia nigra FDR-adjusted p=0.023) and *MBD4* (HR=1.36, 95% CI: 1.12-1.65, Brain – Frontal Cortex BA9 FDR p=0.042) were associated with worse prognoses among TMZ-treated patients in single tissue analyses.

Further subtyping revealed specific associations within *IDH*-mutant and 1p/19q non-codeleted gliomas (*PNKP*: HR=1.65, 95% CI: 1.30-2.10, Brain – Substantia nigra FDR *p*<0.00001 | *MBD4*: HR=1.39, 95% CI: 1.11-1.73, Brain – Frontal Cortex BA9 FDR p=0.078). In this subtype, we observed high concordance and omnibus significant effect of the *PNKP* association across tissues (FDR P_ACAT-O_=0.0147, **Figure 3d**), and altering effects across tissues for *MBD4*. Elevated expression of *HMCES* was associated with improved TMZ-specific *IDH* mutant survival (HR=0.74, 95% CI: 0.61-0.89, Brain – Cerebellum FDR p=0.044), with a similar specific effect in *IDH* mutant 1p/19q non-codeleted gliomas (HR=0.72, 95% CI: 0.58-0.90, Brain – Cerebellum FDR p=0.086).

Specific associations amongst TMZ-treated *IDH*-mutant and 1p/19q codeleted glioma patients included *NEIL2* and *POLG*. Predicted elevated *NEIL2* expression was suggestively associated (FDR *P*_ACAT-O_=0.0873) with decreased survival for TMZ-treated patients across brain tissues. The strongest associations were using basal ganglia-and cortex-specific expression (HR=2.43, 95% CI: 1.44-4.1, Brain – Putamen basal ganglia FDR p=0.015, **Figure 3c**). Elevated *POLG* expression was similarly associated with increased mortality using basal ganglia-specific tissue expression (HR=1.89, 95% CI: 1.22-2.94, Brain – Putamen basal ganglia FDR p=0.037), with direction of effect consistent across 9 of the 13 brain tissues.

In single tissue analyses, increased expression of *TDG*, a base excision repair gene, was associated with increased survival in patients with *IDH*-wildtype glioblastomas treated with TMZ (HR=0.88, 95% CI: 0.82-0.95, Brain – Caudate basal ganglia FDR p=0.029, **Figure 3b**), with a consistent, although not significant (after multiple testing correction) effect across brain tissues (P_ACAT-O_=0.0073).

## Discussion

In the era of precision and genomic medicine, the management of patients with glioma can be done on an individual level basis accounting for the wide breadth of current research and best practices. Temozolomide exists as a crucial resource in the treatment of gliomas, but its effectiveness varies from person to person, and crucially, between glioma subtypes. A growing body of research has shown that *IDH* mutation and 1p/19q chromosomal arm codeletion separately induce higher sensitivity to alkylating chemotherapy, including improved survival, due to changes to the expression of genes controlling DDR pathways^29–31^, highlighting the importance of considering molecular subtyping.

We investigated the association of germline alterations to 34 DNA damage repair genes potentially implicated in the mechanism of action for TMZ and survival time in a cohort of glioma cases spanning the widespread use of TMZ. Using an independent group of glioma cases who were not treated with TMZ, we hoped to identify drug-specific effects. Upon testing 743 putatively functional SNPs we observed statistically significant survival associations for multiple glioma subtypes. Further transcriptome-based analyses revealed broader significant gene-level expression associations. We observed unique associations for differing glioma subtypes, highlighting the distinct biology, microenvironment, and specialized challenges in treating gliomas.

### A germline MGMT variant is associated with glioma-wide survival, possibly through TMZ complications

Our only observation that was significant across glioma subtypes, was the well-studied and common (MAF=0.13 in Europeans) missense variant, rs2308321-G, in *MGMT*, which was associated with decreased survival amongst those treated with TMZ. Previous studies show contradictory associations between this functional SNP with elevated risk of glioma^32^, and decreased risk of glioblastoma^33^. Methylation at the O^6^ position of guanine is a mechanism critical to the anti-tumor activity of TMZ, and *MGMT*’s main role in TMZ resistance is the removal of these damaging lesions. Transfection experiments in e-coli demonstrated the rs2308321-G variant repaired this damage with the same efficiency as the wildtype^34^, suggesting the variant does not alter this mechanism of action. The variant has however been implicated in TMZ myelotoxicity in multiple studies^35–37^. Though we lack information on TMZ related toxicities in our cohort, a possible explanation for the effect of rs2308321-G on survival could be that it leads to lower exposure to TMZ from dose reductions, delays, and/or premature discontinuation as a result of myelosuppression.

### Germline markers in IDH-mutant gliomas may increase chemosensitivity through multiple pathways

Temozolomide resistance and development of hypermutation at recurrence is a serious complication amongst individuals with *IDH*-mutant glioma. The presence of *MGMT* promoter methylation levels at diagnosis is the best established marker for risk^10^. Generally, resistance to therapeutics continues to be a major cause of treatment failure for individuals with cancer. Anticipating how germline differences can alter response to TMZ, particularly in light of known alternative chemotherapy options, may influence treatment decisions for certain patients. The recently completed promising clinical trials of vorasidenib, a pan mutant *IDH* inhibitor, presents one possible novel treatment avenue for *IDH*-mutant gliomas^38^.

Our single variant analysis of *IDH*-mutant gliomas identified a highly pathogenic variant, rs13076508-C located near *PARP3*, was associated with improved survival in those treated with TMZ. This variant was associated with altered expression and splicing variants of multiple nearby genes within the brain. Amongst those genes, this variant was associated with significant downregulation of toll-like receptor 9 (*TLR9)* and BRCA-1 Associated Protein-1 (*BAP1)* genes within the brain. *TLR9* is one of 10 toll-like receptor genes, which are generally expressed by antigen presenting cells, and play key roles in adaptive immunity. *TLR9* specifically plays the essential role of recognizing patterns of unmethylated CpG DNA in order to identify foreign versus self-antigens in innate immune activation^39^. Overexpression of *TLR9* was a negative prognostic factor in many cancers, including glioma^40–42^10/10/2024 11:18:00 AM, although the opposite was observed in pancreatic and breast cancers . TLR agonists were previously shown to sensitize cells to chemotherapy, and used in several recent clinical trials evaluating their safety and effectiveness in combination with existing therapies across multiple cancers^45^.

Our variant of interest was also associated with significantly decreased *BAP1* expression in the brain. *BAP1* plays many roles in maintaining genomic stability and was recently identified to be recruited by *PARP1* during DNA damage repair^46^. *PARP1* is a major component of the base excision repair pathway, and its inhibition has shown to sensitize glioma cell lines to TMZ and decrease drug resistance, independent of *MGMT* promoter methylation status^47^. This line of evidence suggests that the natural inhibition of *BAP1* (through germline variation) may confer tumor sensitization to TMZ. It is unclear whether the functional mechanism underlying the rs13076508-C association acts through *TLR9, BAP1*, or a combination of both, but suggests fruitful directions for further research.

### Decreased PNKP expression may solely benefit IDH-mutant gliomas without 19q arm deletion

We identified a strong association between *PNKP* expression and survival in glioma cases with *IDH* mutation but without 1p/19q codeletion that were treated with TMZ. We observed that genetically predicted low transcript levels of *PNKP* within the brain were associated with significantly longer survival compared to those with predicted elevated/normal levels. This association suggests germline inhibition of the PNKP gene may offer a natural increased chemosensitivity. Previous functional work identified a similar effect, that a knockdown of *PNKP* expression using siRNAs resulted in increased TMZ sensitivity *in vitro* using glioma cell lines, with an approximate 50% reduction in surviving cells compared to the unaltered control^48^. *PNKP* is located on the q arm of chromosome 19, as such its expression is likely altered between the two distinct *IDH* mutant subtypes, aligning with previous observations that the somatic loss of heterozygosity of the 19q arm was associated with improved chemosensitivity^29^. Expectedly, we saw no association with *PNKP* within our 1p/19q codeleted cases. *PNKP* is highly linked with fellow BER gene, *XRCC1*, in the process of DNA damage repair^49^, and studies of their interaction could help identify possible mechanism. Further study into *PNKP* expression in *IDH*-mutant 1p/19q non-codeleted gliomas is warranted.

### NEIL2 markers may assist in treatment choices for patients with IDH-mutant 1p/19q codeleted gliomas

In the treatment of *IDH*-mutant and 1p/19q codeleted gliomas, there is controversy regarding whether either TMZ or procarbazine, lomustine, and vincristine (PCV) in combination with radiation therapy leads to better outcomes, with the ongoing CODEL clinical trial investigating the impact on progression-free survival for each drug^50^. In our analyses, germline alterations linked to depleted *NEIL2* expression were associated with improved survival for patients with *IDH*-mutant 1p/19q codel gliomas treated with TMZ, suggesting an increased chemosensitivity. Overabundance of the *NEIL2* protein has been associated with decreased sensitivity to other chemotherapeutics^51^ consistent with our findings. Further study to replicate these findings, including if they extend to PCV treatment, and to understand the underlying mechanism are warranted.

### Two markers may be linked to increased intrinsic chemo-sensitivity in IDH-wildtype glioblastomas

The role of TMZ in treating *IDH*-wildtype glioblastomas is well-understood, with a clear improvement in survival when combined with other standard-of-care treatments such as radiation and maximum safe surgical resection, particularly in patients with *MGMT* promoter methylation^4^. While it remains standard of care, intrinsic TMZ resistance does occur in a number of glioblastoma patients, and while strategies to sensitize cells have been explored, including *STAT3* and *MGMT* inhibitors^52^, there has been little observed benefit. The question remains if TMZ should be administered once evidence of its ineffectiveness exists, particularly due to the short survival time of glioblastoma patients.

Our study nominates two possible markers, outside of *MGMT* promoter methylation, that may be useful in identifying TMZ effectiveness in *IDH*-wildtype glioblastoma patients prior to drug administration. We first identified a rare pathogenic SNP near the uracil DNA glycosylase (*UNG*) gene on chromosome 12. Cell line knockdowns of UNG demonstrated increased TMZ sensitivity, with mechanism of action hypothesized to be through UNG’s ability to repair lethal oxidative lesions formed from an increase in reactive oxygen species initiated through high dose TMZ^53^. Despite our work demonstrating a 3 month decrease in TMZ-related survival for those carrying the rare variant, we were unable to find any functional association with this SNP. Further research and replication are warranted to validate this marker. We observed a brain-tissue specific association that genetically predicted Thymine DNA glycosylase (*TDG)* overexpression was linked to increased glioblastoma survival, given TMZ treatment. We note that this association, however, was not significant in our omnibus test. *TDG* is involved in active DNA demethylation and DNA damage repair and is highly expressed in gliobastomas^54^. A knockout of *TDG* in both human and mouse cells was shown to be sufficient to generate resistance to the common chemotherapeutic drug 5-Fluoroacil (5-FU), and that overexpression increased sensitivity^55^, consistent with our observed direction of effect with TMZ. Interestingly, the two nominated genes, *UNG* and *TDG*, have highly overlapping function in DNA repair^56^. *UNG* and *TDG* expression vary widely within GBMs^53^, and studies integrating TMZ usage, tumor expression, and germline mutations may help disentangle their potential effects on drug sensitivity.

### Strengths and weaknesses of the presented study

Our findings should still be considered with limitations. This study was based on a pair of historic cohorts. As such, we lacked the full suite of somatic molecular markers to adhere to strict WHO 2021 classification guidelines. *MGMT* promoter methylation status, a relatively recently identified and costly phenotype to measure, is unknown in our cohort, and so we were unable to consider it in our analyses. Although overall survival times remain similar, treatments have advanced and patients overall are showing slightly increased survival, which makes comparing survival across historic cohort more challenging. We do overcome this by not directly comparing the survival of patients between the pre- and post-TMZ eras. The datasets do not include robust information on the recurrent tumor, including possible TMZ-induced hypermutation. Our results may not extend broadly extend to all ancestries, as the set of available patients were overwhelmingly of European descent. Our definition of TMZ treatment for the purposes of this analysis was limited to a binary yes/no history of known drug usage during the first course of treatment. Ongoing studies which collect detailed treatment information across the course of the disease will be key to determining if the effects presented in this study are indeed isolated to just first-course treatment.

Despite these limitations, this analysis is one of the most extensive studies of germline genetic effects on TMZ, through the survival of patients with glioma with key molecular subtyping and long-term follow-up. The strongest aspect of this study is the decades’ long recruitment periods of cases spanning more than 20 years, with substantial recruitment in both the pre-TMZ and post-TMZ eras. This created a unique opportunity to study effects of the drug with a truly naïve comparison group, which has become a challenge for modern cohorts as TMZ has now become standard of care.

## Conclusions

This study found statistically significant associations of germline variants in DNA damage repair pathways potentially mechanistically linked to TMZ action among glioma patients who received TMZ as part of first-course treatment but not in those who were not initially treated with TMZ. Overall, our results are consistent with previous literature on DNA damage repair genes and chemotherapy sensitization. While further validation is needed, this study lays strong groundwork for the concept of variation in the intrinsic sensitivity to TMZ. The markers identified can help to identify possible targets to complement TMZ to improve accurate clinical management and glioma patient survival.

## Data Availability

Genotype data of glioma samples from Mayo Clinic and control samples from the Glioma International Case Control Study (GICC) are available from dbGaP under accession phs001319.v1.p1. Genotype data from the University of California, San Francisco Adult Glioma Study (AGS) are available under dbGap accession phs001497.v2.p1.

## List of abbreviations

IDH: Isocitrate dehydrogenase 1 and 2 genes
1p/19q: 1p and 19q chromosomal arms
TMZ: Temozolomide
DDR: DNA damage repair
BER: Base excision repair
MMR: Mismatch repair
UCSF: University of California San Francisco
AGS: Adult Glioma Study
FDR: False discovery rate
HR: Hazard ratio
PH: Proportional Hazards
KM: Kaplan-Meier
CI: Confidence interval
WHO: World Health Organization
TCGA: The Cancer Genome Atlas
GTEx: Genotype-Tissue Expression Project
eQTL: Expression Quantitative Trait Loci
sQTL: Splicing Quantitive Trait Loci

## Declarations

### Ethics approval and consent to participate

Collection of patient samples and associated clinicopathological information was undertaken with written informed consent and relevant ethical review board approval at the respective study centers in accordance with the tenets of the Declaration of Helsinki. Specifically informed consent and ethical board approval was obtained from the UCSF Committee on Human Research (USA) and the Mayo Clinic Office for Human Research Protection (USA). The diagnosis of glioma (ICDO-3 codes 9380-9480 or equivalent) was established through histology and somatic molecular markers in all cases in accordance with World Health Organization guidelines.

## Funding

Work at the University of California, San Francisco, was supported by the National Institutes of Health (grant numbers T32CA112355, R01CA52689, P50CA097257, R01CA126831, R01CA139020, and R01CA266676), as well as the loglio Collective, the National Brain Tumor Foundation, the Stanley D. Lewis and Virginia S. Lewis Endowed Chair in Brain Tumor Research, the Robert Magnin Newman Endowed Chair in Neuro-oncology, and by donations from families and friends of John Berardi, Helen Glaser, Elvera Olsen, Raymond E. Cooper, and William Martinusen.

The work at Mayo was supported by National Cancer Institute (NCI) grants CA230712, P50CA108961, and CA139020; the National Brain Tumor Society; the loglio Collective; the Mayo Clinic; and the Ting Tsung and Wei Fong Chao Foundation.

The work at Stanford University was supported by the National Institutes of Health grant R00CA246076.

This publication was supported by the National Center for Research Resources and the National Center for Advancing Translational Sciences, National Institutes of Health, through UCSF-CTSI Grant Number UL1 RR024131. Its contents are solely the authors’ responsibility and do not necessarily represent the official views of the NIH. The authors wish to acknowledge study participants, the clinicians, and the research staff at the participating medical centers, the UCSF Cancer Registry, and the UCSF Neurosurgery Tissue Bank.

The collection of cancer incidence data used in this study was supported by the California Department of Public Health pursuant to California Health and Safety Code Section 103885; Centers for Disease Control and Prevention’s (CDC) National Program of Cancer Registries, under cooperative agreement 5NU58DP006344; the National Cancer Institute’s Surveillance, Epidemiology and End Results Program under contract HHSN261201800032I awarded to the University of California, San Francisco, contract HHSN261201800015I awarded to the University of Southern California, and contract HHSN261201800009I awarded to the Public Health Institute, Cancer Registry of Greater California. The ideas and opinions expressed herein are those of the author(s) and do not necessarily reflect the opinions of the State of California, Department of Public Health, the National Cancer Institute, and the Centers for Disease Control and Prevention or their Contractors and

Subcontractors. All analyses, interpretations, and conclusions reached in this manuscript from the mortality data are those of the author(s) and not the State of California Department of Public Health.

## Authorship

GG, VG, LC, AH, VK, JS, and SSF conceived of the study. GG and SSF wrote the main drafts of the manuscript. GG, GW, and SSF conducted statistical and computational analyses. GG, SSF, JWT, JLC, AMM, JLW, JEP, RBJ, LK, ERA, and MW advised on result interpretations. The primary data collection involved LM, HMH, AMM, JLW, TR, JEP, RBJ, JKW, and MW. All authors read and approved the final manuscript.

## Acknowledgements

The Genotype-Tissue Expression (GTEx) Project was supported by the <u>Common Fund</u> of the Office of the Director of the National Institutes of Health, and by NCI, NHGRI, NHLBI, NIDA, NIMH, and NINDS. The data used for the analyses described in this manuscript were obtained from the GTEx Portal on 04/26/23.

The results published here are in whole or part based upon data generated by The Cancer Genome Atlas managed by the NCI and NHGRI. Information about TCGA can be found at http://cancergenome.nih.gov.

