## Supplemental Figures and Tables for "Functional germline variants in DNA damage repair pathways are associated with altered survival in adults with glioma treated with temozolomide"

**Supplemental Table S1: Clinical and molecular summary of included adults with IDH specific gliomas with known usage/absence of temozolomide treatment.**

|  | **IDH mutant** | | **IDH wildtype** | |
| --- | --- | --- | --- | --- |
|  | **Received TMZ** | **No TMZ** | **Received TMZ** | **No TMZ** |
| Number of cases (%) | 260 (54.97%) | 213 (45.03%) | 780 (75.58%) | 252 (24.42%) |
| Age at diagnosis (Median + IQR) | 40 (IQR: 16) | 39 (IQR: 17) | 58 (IQR: 15) | 58 (IQR: 18) |
| Sex (% Male) | 61.15% | 60.38% | 64.49% | 57.14% |
| Genotyping array |  |  |  |  |
| Oncoarray | 76.54% | 82.55% | 65.77% | 42.46% |
| i370 | 23.46% | 17.45% | 34.23% | 57.54% |
| Data source |  |  |  |  |
| UCSF AGS | 79.62% | 69.81% | 76.15% | 74.21% |
| Mayo Clinic | 20.38% | 30.19% | 23.85% | 25.79% |
| 1p/19q |  |  |  |  |
| Codeleted | 35.00% | 36.32% | 86.79% | 85.32% |
| Non-codeleted | 65.00% | 63.68% | 0.26% | 1.59% |
| NA | 0% | 0% | 12.95% | 13.10% |
| *TERT* |  |  |  |  |
| Wildtype | 51.54% | 51.42% | 14.74% | 16.27% |
| Mutant | 35.00% | 29.72% | 67.82% | 63.10% |
| NA | 13.46% | 18.87% | 17.44% | 20.63% |
| Grade (% of total) |  |  |  |  |
| 2 | 37.69% | 65.57% | 3.08% | 13.10% |
| 3 | 43.08% | 25.47% | 11.41% | 13.89% |
| 4 | 19.23% | 8.96% | 85.51% | 73.02% |
| Histology* (% of total) |  |  |  |  |
| Astrocytoma | 34.62% | 33.96% | 13.08% | 16.27% |
| Oligodendroglioma | 28.08% | 33.02% | 0.64% | 6.35% |
| Glioblastoma | 18.85% | 8.02% | 85.13% | 73.02% |
| Oligoastrocytoma | 18.46% | 25.00% | 1.15% | 4.37% |
| Diagnosis Year  Median (min, max, IQR) | 2007 (‘98, ‘13, 5) | 2003 (‘91, ‘13, 10) | 2008 (‘01, ‘14, 6) | 2002 (‘91, ‘14, 9) |
| % Censored (no known death event) | 46.92% | 42.45% | 6.15% | 8.33% |
| Chemotherapy (% known Chemo) | 100% | 27.83% | 100% | 40.87% |
| Radiation (% known RT) | 70.38% | 51.42% | 97.44% | 87.70% |

*As classified using World Health Organization 2016 central nervous system classification guidelines.

**Supplemental Table S2: Tissue-specific genetically imputed transcript levels of DNA repair genes suggestively associated with survival amongst glioma cases treated with temozolomide.**

|  | **Gene** | **Chr** | **Tissue Source** | **Tissue Site** | **HR (95% CI)** | **Cox P-value (FDR adjusted)** |
| --- | --- | --- | --- | --- | --- | --- |
| ***All glioma*** | | | | | | |
|  | *TDG* | 12 | GTEx | Brain Caudate basal ganglia | 0.90 (0.84-0.96) | 0.062 |
|  | *OGG1* | 3 | GTEx | Whole blood | 1.11 (1.04-1.19) | 0.076 |
| ***IDH* mutant 1p/19q codeleted glioma** | | | | | | |
|  | *MPG* | 16 | GTEx | Brain Amygdala | 0.57 (0.37-0.89) | 0.10 |
|  | *MUTYH* | 1 | TCGA | LGG Tumor | 1.79 (1.12-2.86) | 0.064 |
|  | *NEIL2* | 8 | GTEx | Brain Cortex | 2.04 (1.27-3.26) | 0.052 |
|  | *NEIL2* | 8 | TCGA | LGG Tumor | 1.81 (1.14-2.88) | 0.064 |
|  | *PARP3* | 3 | GTEx | Brain Frontal cortex | 1.98 (1.21-3.24) | 0.096 |
|  | *SMUG1* | 12 | GTEx | Brain Amygdala | 0.44 (0.24-0.78) | 0.084 |
|  | *UNG* | 12 | TCGA | LGG Tumor | 0.55 (0.33-0.89) | 0.064 |
| ***IDH* mutant 1p/1q non-codeleted glioma** | | | | | | |
|  | *HMCES* | 3 | GTEx | Brain Cerebellum | 0.72 (0.58-0.90) | 0.086 |
|  | *MBD4* | 3 | GTEx | Brain Frontal cortex | 1.39 (1.11-1.73) | 0.078 |
